# Sodium content of menu items in New York City chain restaurants following enforcement of the sodium warning icon rule, 2015-2017

**DOI:** 10.1101/2022.09.02.22279522

**Authors:** Julia S. Sisti, Divya Prasad, Sarah Niederman, Tamar Adjoian Mezzacca, Amaka V. Anekwe, Jenifer Clapp, Shannon M. Farley

**Affiliations:** Bureau of Chronic Disease Prevention, New York City Department of Health and Mental Hygiene

## Abstract

In 2016, New York City (NYC) began enforcing a sodium warning regulation at chain restaurants, requiring placement of an icon next to any menu item containing ≥2,300 mg sodium. As menu labeling may improve menu nutritional composition, we investigated whether sodium content of menu items changed following enforcement of the sodium warning icon. All menu offerings at 10 quick-service (QSR) and 3 full-service (FSR) chain restaurants were photographed in 2015 (baseline) and 2017 (follow-up) and matched to nutritional information from restaurant websites; items were categorized as being available at both baseline and follow-up, or at only one timepoint. Linear and logistic regression models, respectively, assessed changes in calculated mean sodium-per-serving and the odds of an item containing ≥2,300 mg sodium. At baseline, mean per-serving sodium content was 2,160 mg at FSR and 1,070 mg at QSR, and 40.6% of FSR items and 7.2% of QSR items contained ≥2,300 mg sodium per serving. Sodium content did not differ when comparing all items offered at follow-up to all offered at baseline (21 mg, 95% CI: -60,101), or when comparing new versus discontinued items (17 mg, 95% CI: -154, 187). At follow-up, there was a non-significant increase in the overall likelihood of items requiring a warning icon (OR=1.32, 95% CI: 0.97,1.79). When comparing new versus discontinued items, there was a twofold increase in the odds of requiring a warning icon (OR=2.08, 95% CI: 1.02,4.24). Our findings both highlight high sodium content of menu items at popular chain restaurants and underscore difficulties in motivating restaurants to reduce sodium levels.

## Introduction

High sodium intake is a well-established, modifiable risk factor for hypertension [1] and a leading dietary contributor to cardiovascular mortality [2-3]. Food purchased outside the home accounts for over two-thirds of dietary sodium intake [4-5], and frequency of dining out is positively associated with high sodium intake among Americans [6]. In New York City (NYC), average daily sodium intake is estimated to be about 3,200 milligrams (mg) per day [7], approximately 40% higher than the daily recommended sodium limit for healthy adults (2,300 mg) [8].

In December 2015, in light of this landscape and in an effort to foster greater transparency of excessive sodium in the food supply, NYC enacted regulation requiring chain restaurants to post a sodium warning icon depicting a saltshaker next to any menu item containing ≥2,300 mg sodium [9]. In addition to standard menu items, customizable items require a warning icon if any potential combination of components contains ≥2,300 mg sodium; this also applies to combination meals with multiple components sold together at a fixed price. Shareable items containing ≥2,300 mg sodium per serving must also display an icon. Warning icons must appear on all printed or electronic menus, menu boards and item tags; additionally, restaurants must display a statement at the point of purchase which explains the meaning of the icon and summarizes the association of high sodium intake with heart disease and stroke.

In addition to informing purchasing decisions, menu labeling at chain restaurants may lead to improvements in the nutritional composition of offered menu items. In an analysis of 59 national chain restaurants between 2012-2019, Grummon *et al*. reported that menu items newly introduced following implementation of calorie labeling, in accordance with the federal requirement, were lower in calories than items newly introduced prior to calorie labeling [10]. We investigated whether the sodium content of menu items offered in NYC chain restaurants changed following enforcement of the sodium warning rule in 2016.

## Methods

### Study population

The NYC Department of Health and Mental Hygiene (Health Department) maintains a list of approximately 27,000 current restaurants licensed by the city; those with ≥15 locations nationwide are considered chains and covered by the sodium warning icon rule. In 2015, a total of 182 NYC restaurant chains, representing 2,129 locations, were required to comply with the rule. Of these, 50 chains had table service and were defined as ‘full-service restaurants’ (FSR); the remainder were classified as ‘quick service restaurants (QSR)’. We identified the 10 QSR and 5 FSR chains with the greatest number of NYC locations in 2015; of these, two FSR were missing sodium data for at least one study time point and were excluded from analyses. No human subjects were involved in this research and therefore IRB review was not required.

### Data collection

Baseline data collection occurred between November 2015 and January 2016, prior to enforcement of the warning icon rule by the Health Department in June 2016; follow-up data collection occurred in March-April 2017. Trained data collectors visited a single NYC location of each chain and photographed all printed menus, menu boards, signage, and display cases. At each time point, images were matched with nutritional information on restaurant websites. When possible, the same location was visited at both time points, to reduce potential menu variation across different locations of a single chain.

### Measures

We calculated per-serving sodium content of menu items offered in each year. To reflect rule enforcement, each customizable and combination meal was included as a single item, with the highest sodium components included. When customizable options included different protein bases (e.g. chicken, pork, beef), we created separate items for each protein that additionally included the highest sodium options for all other components. For customizable pizzas, we created separate items for each crust option (i.e. thin crust, regular) that included the three highest sodium toppings. Different sizes of single items (e.g., small, medium and large fries) were entered as separate items. When sodium content for an item was presented as a range, we used the maximum value. Sodium content was missing for a large proportion of beverages (>35%); therefore, beverages were not examined individually but were included in combination meals when sodium content was available. We additionally excluded items described as ‘shareable’, and for which per-serving sodium content could not be calculated. Toppings, dressings, and dipping sauces were not analyzed except as part of customizable meals, as they could not be ordered individually and thus were not eligible for a warning icon. Sodium information was collected for a small number of children’s menu items (33 at baseline; 35 at follow-up); exclusion of these items did not change results and they were retained in final analyses.

Menu items were classified as combination meals if they included two or more à la carte items (e.g., a hamburger and fries), offered together at a fixed price. Side dishes included fries, appetizers, salads and soups described as sides, baked goods, and desserts; main dishes included entrées, pizza, sandwiches, hamburgers, and large salads and soups.

### Statistical analysis

We used two approaches to evaluate changes in sodium content between time points. First, to test for differences in mean sodium content, we used linear regression models with per-serving sodium content (mg) as the outcome. Second, logistic regression models assessed whether the odds of an item containing ≥2,300 mg sodium per serving differed between baseline and follow-up. Models included a dichotomous indicator for time point, and adjusted for item type (combination meal, main dish, side dish) and restaurant type (QSR, FSR). To account for correlation of sodium content within chains, models included random intercepts for restaurant chain.

In addition to comparing all items offered at baseline to all items offered at follow-up, we separately compared 1) items that were available at both baseline and follow-up; and 2) items that were available at follow-up only versus those available at baseline only. Items that were offered at both time points were included if sodium information was available at either point; results were unchanged when restricting to those with complete data at both points.

## Results

Our analysis includes 10 QSR chains (Baskin Robbins, Burger King, Chipotle, Domino’s, Dunkin’ Donuts, McDonald’s, Papa John’s, Popeyes, Starbucks, Subway) with a total of 2,045 NYC locations and three FSR chains (Applebee’s, IHOP, TGI Friday’s) with a total of 57 NYC locations, accounting for 64% and 24% of all NYC QSR and FSR chain locations, respectively. Mean time between baseline and follow-up data collection was 15.6 months. At baseline, one FSR chain (Applebee’s) had voluntarily added the warning icon in advance of enforcement. Analyses include a total of 840 menu items available at baseline and 887 items offered at follow-up; descriptive statistics for items are shown in Table 1. At baseline, mean per-serving sodium content was twofold higher among items offered at FSR (2,160 mg) compared to those at QSR (1,070 mg). At baseline, 40.6% of FSR menu items had ≥2,300 mg sodium per serving, compared to only 7.2% of QSR menu items (Table 2); five QSR had no items exceeding this threshold at either time point.

**Table 1.** Mean per-serving sodium content (mg) of food items offered at 13 NYC chain restaurants at baseline and follow-up.

|  |  | All Y1 items vs.<br>all Y2 items |  |  | Items offered in both<br>Y1 and Y2 |  |  | Items offered in Y1 only vs.<br>Y2 only |  |  |
| --- | --- | --- | --- | --- | --- | --- | --- | --- | --- | --- |
|  |  | n (Y1/Y2) | Y1 | Y2 | n (Y1/Y2) | Y1 | Y2 | n (Y1/Y2) | Y1 | Y2 |
| **Overall** |  | **840/887** | **1444** | **1406** | **655/681** | **1414** | **1393** | **185/206** | **1550** | **1451** |
|  | Main dish | 340/362 | 1402 | 1372 | 268/279 | 1409 | 1393 | 72/83 | 1378 | 1299 |
|  | Side dish | 251/274 | 671 | 644 | 209/223 | 669 | 672 | 42/51 | 681 | 525 |
|  | Combination meal | 249/251 | 2280 | 2288 | 178/179 | 2296 | 2290 | 71/72 | 2237 | 2281 |
| **Quick service** |  | **552/612** | **1070** | **1023** | **437/464** | **1037** | **1020** | **115/148** | **1196** | **1034** |
|  | Main dish | 226/261 | 1160 | 1124 | 183/195 | 1160 | 1141 | 43/66 | 1159 | 1073 |
|  | Side dish | 182/212 | 412 | 404 | 157/171 | 415 | 432 | 25/41 | 392 | 287 |
|  | Combination meal | 144/139 | 1762 | 1778 | 97/98 | 1812 | 1803 | 47/41 | 1659 | 1719 |
| **Full service** |  | **288/275** | **2160** | **2258** | **218/217** | **2170** | **2190** | **70/58** | **2130** | **2514** |
|  | Main dish | 114/101 | 1883 | 2012 | 85/84 | 1944 | 1979 | 29/17 | 1703 | 2176 |
|  | Side dish | 69/62 | 1357 | 1465 | 52/52 | 1439 | 1458 | 17/10 | 1108 | 1502 |
|  | Combination meal | 105/112 | 2989 | 2920 | 81/81 | 2876 | 2879 | 24/31 | 3371 | 3025 |

**Table 2.** Proportion of food items containing ≥2,300 mg sodium/serving at 13 NYC chain restaurants at baseline and follow-up.

|  | All Y1 items vs.<br>all Y2 items |  |  | Items offered in both<br>Y1 and Y2 |  |  | Items offered in Y1 only vs.<br>Y2 only |  |  |
| --- | --- | --- | --- | --- | --- | --- | --- | --- | --- |
|  | n (Y1/Y2) | Y1 | Y2 | n (Y1/Y2) | Y1 | Y2 | n (Y1/Y2) | Y1 | Y2 |
| **Overall** | **840/887** | **18.7** | **20.5** | **655/681** | **19.2** | **19.8** | **185/206** | **16.8** | **22.8** |
| Main dish | 340/362 | 14.4 | 15.5 | 268/279 | 14.9 | 16.1 | 72/83 | 12.5 | 13.3 |
| Side dish | 251/274 | 5.2 | 5.1 | 209/223 | 5.7 | 5.4 | 42/51 | 2.4 | 3.9 |
| Combination meal | 249/251 | 38.2 | 44.6 | 178/179 | 41.6 | 43.6 | 71/72 | 29.6 | 47.2 |
| **Quick service** | **552/612** | **7.2** | **7.7** | **437/464** | **7.8** | **7.5** | **115/148** | **5.2** | **8.1** |
| Main dish | 226/261 | 8.0 | 7.3 | 183/195 | 8.2 | 8.2 | 43/66 | 7.0 | 4.5 |
| Side dish | 182/212 | 0 | 0 | 157/171 | 0 | 0 | 25/41 | 0 | 0 |
| Combination meal | 144/139 | 15.3 | 20.1 | 97/98 | 20.0 | 19.4 | 47/41 | 6.4 | 22.0 |
| **Full service** | **288/275** | **40.6** | **49.1** | **218/217** | **42.2** | **46.1** | **70/58** | **35.7** | **60.3** |
| Main dish | 114/101 | 27.2 | 36.6 | 85/84 | 29.4 | 34.5 | 29/17 | 20.7 | 47.1 |
| Side dish | 69/62 | 18.8 | 22.6 | 52/52 | 23.1 | 23.1 | 17/10 | 5.9 | 20.0 |
| Combination meal | 105/112 | 69.5 | 75.0 | 81/81 | 67.9 | 72.8 | 24/31 | 75.0 | 80.6 |

Table 3 presents unadjusted and multivariable-adjusted differences and 95% confidence intervals (95% CI) in mean per-serving sodium content of menu items between time points. No significant difference in sodium content was observed when comparing all items offered at follow-up to all offered at baseline (21mg, 95% CI=-60, 101). Similarly, no differences between follow-up and baseline sodium content were observed among items offered in both years (14mg, 95% CI=-79, 107) or comparing newly introduced versus discontinued items (17mg, 95% CI=-154, 187).

**Table 3.**
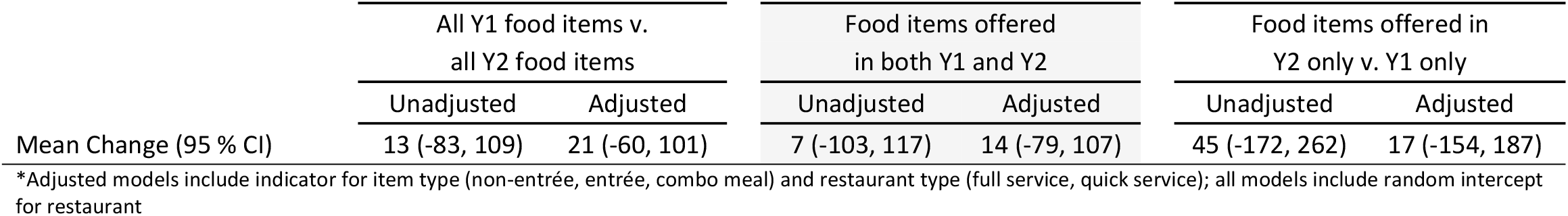
Mean per-serving change in sodium content (mg) of food items comparing Y2 (follow up) vs. Y1 (baseline)*

Odds ratios (OR) comparing the likelihood of an item meeting or exceeding the 2,300 mg sodium threshold are shown in Table 4. Compared to baseline, there was a non-significant increase in the overall likelihood of menu items at follow-up requiring a warning icon (OR=1.32, 95% CI=0.97, 1.79). When comparing new items to those that had been discontinued, there was a statistically significant twofold increase in odds of requiring a warning icon (OR=2.08, 95% CI=1.02, 4.24); conversely, no significant increase was observed when comparing items available in both time points (OR=1.15, 95% CI=0.82, 1.63).

**Table 4.** Odds of food item requiring sodium warning icon (≥2,300 mg sodium/serving), comparing Y2 (follow-up) vs. Y1 (baseline)*

|  | All Y1 food items v.<br>all Y2 food items |  | Food items offered<br>in both Y1 and Y2 |  | Food items offered in<br>Y2 only v. Y1 only |  |
| --- | --- | --- | --- | --- | --- | --- |
|  | Unadjusted | Adjusted | Unadjusted | Adjusted | Unadjusted | Adjusted |
| Odds Ratio (95% CI) | 1.30 (0.99, 1.72) | 1.32 (0.97, 1.79) | 1.12 (0.82, 1.52) | 1.15 (0.82, 1.63) | 2.12 (1.14, 3.96) | 2.08 (1.02, 4.24) |

\*Adjusted models include indicator for item type (non-entrée, entrée, combo meal) and restaurant type (full service, quick service); all models include random intercept for restaurant

## Discussion

In our analysis of 13 NYC chain restaurants, we observed no evidence that overall sodium content of menu items changed immediately following enforcement of the sodium warning rule; however, we found that newly introduced items were more likely to contain at least 2,300 mg of sodium and require a sodium warning icon compared to those that had been discontinued. Notably, sodium content of menu items was high in both time periods, particularly at FSR, where nearly half of menu items required a warning icon.

To our knowledge, only one previous analysis has examined the effect of mandatory restaurant menu labeling on sodium content [11]. Eighteen months after King County, Washington required chain restaurants to provide calorie, sodium and saturated fat content for menu items in 2009, sodium content was modestly lower among entrées served at sit-down, but not quick-service, restaurants. However, the WA law only required restaurants to provide sodium content (in mg) at point of purchase, including in brochures or menu appendices. Additionally, baseline for this analysis was 6 months post-implementation of regulations; pre-implementation sodium levels were not examined.

Overall trends in sodium content of restaurant food were examined by Wolfson *et al*. [12], in an analysis of menu items at 66 large chain restaurants from 2012-2016. Decreased levels of sodium were observed when comparing items added in subsequent years to those available in 2012 only. No trends were observed for items offered in all study years, suggesting that rather than reformulate existing menu items, restaurants may be introducing new, lower-sodium options in addition to their standard offerings, a hypothesis supported by at least one other analysis of 213 restaurants [13]. Other studies have included fewer restaurants; some [14-15], but not all [16], have shown increases in sodium content over time.

A primary strength of our study is our in-store data collection, which allowed us to account for potential regional variation in menu offerings within a single chain, and to compute sodium content for customizable and combination meals, which appear frequently on menus and routinely exceed recommended sodium limits. However, limitations of our analysis must be acknowledged. First, we examined a relatively small sample of restaurants, including several of the largest national chains, which may be less likely than smaller chains to decrease sodium content in response to local regulations. However, in selecting chains that account for a large proportion of NYC chain restaurant locations, we were able examine those menu items that were most readily available to consumers. Second, because we calculated sodium content of customizable meals by summing the highest-sodium components, our estimates for these meal types only reflect changes to the highest-sodium containing versions and do not reflect the full scope of menu changes, for example the introduction of lower-sodium customizations (i.e. a choice of apple slices or French fries as a side). This methodology was selected to mirror rule enforcement and to assess whether restaurants would change their menus with the aim of reducing the number of items requiring a warning icon, but may restrict comparability with other published studies. Lastly, our follow-up window was relatively short; it is possible that reductions in sodium may occur over a longer period.

Our findings highlight the high sodium content of menu items at popular chain restaurants; they also underscore the difficulty of motivating restaurants to reduce sodium levels, though other efforts are underway. In addition to enacting the sodium warning rule, the Health Department has also played a key role in the National Salt and Sugar Reduction initiative (NSSRI), a coalition of health organizations and state and local health authorities which partnered with the food industry to reduce sodium content in packaged and restaurant food [17]. While the restaurant industry’s progress toward NSSRI targets has not yet been assessed, some improvements in packaged foods have been noted [18]. Further, in October 2021, in recognition that most dietary sodium comes from processed, packaged, and prepared foods, the FDA released voluntary sodium targets to encourage industry, including restaurants, to reduce sodium across a wide range of their products. Our results, combined with those from NSSRI, suggest that current approaches may need to be combined with consumer education and further engagement with industry to produce meaningful reductions in sodium intake and improvements in health outcomes.

## Data Availability

Datafiles and code will be made available on GitHub following manuscript acceptance.

## Acknowledgements

The authors would like to thank John Jasek for thoughtful review and revision of the manuscript.

